# HOW, WHY AND UNDER WHAT CIRCUMSTANCES DOES A QUALITY IMPROVEMENT COLLABORATIVE BUILD KNOWLEDGE AND SKILLS IN CLINICIANS WORKING WITH PEOPLE WITH DEMENTIA? A REALIST INFORMED PROCESS EVALUATION

**DOI:** 10.1101/2020.08.05.20169185

**Authors:** Lenore de la Perrelle, Monica Cations, Gaery Barbery, Gorjana Radisic, Billingsley Kaambwa, Maria Crotty, Janna Anneke Fitzgerald, Susan Kurrle, Ian D Cameron, Craig Whitehead, Jane Thompson, Kate Laver

## Abstract

In increasingly constrained health and aged care services, strategies are needed to improve quality and translate evidence into practice. In dementia care, recent failures in quality and safety have led the World Health Organisation to prioritise the translation of known evidence into practice. While quality improvement collaboratives have been widely used in healthcare, there are few examples in dementia care.

We describe a recent quality improvement collaborative to improve dementia care across Australia and assess the implementation outcomes of acceptability and feasibility of this strategy to translate known evidence into practice. A realist-informed process evaluation was used to analyse how, why and under what circumstances a quality improvement collaborative built knowledge and skills in clinicians working in dementia care.

This realist-informed process evaluation developed, tested, and refined the program theory of a quality improvement collaborative. Data were collected pre-and post-intervention using surveys and interviews with participants (n=24). A combined inductive and deductive data analysis process integrated three frameworks to examine the context and mechanisms of knowledge and skill building in participant clinicians.

A refined program theory showed how and why clinicians built knowledge and skills in quality improvement in dementia care. Seven mechanisms were identified: motivation, accountability, identity, collective learning, credibility, and reflective practice. Each of these mechanisms operated differently according to context.

A quality improvement collaborative designed for clinicians in different contexts and roles was acceptable and feasible in building knowledge and skills of clinicians to improve dementia care. A supportive setting and a credible, flexible, and collaborative process optimises quality improvement knowledge and skills in clinicians working with people with dementia.

**Trial registration:** Australian and New Zealand Clinical Trials Registry 21 February 2018 (ACTRN 12618000268246)

## Background

The challenge of implementing evidence-based guidelines into clinical practice continues to be of concern in healthcare (1). In dementia care recent OECD reports show poor care and low training persists in member countries (2). In Australia serious failures in dementia care (3) have prompted inquiries into safety and quality (4). In this context, the World Health Organisation Global action plan on the public health response to dementia 2017-2025 (5) identified as a priority, the need to translate what is already known about dementia care into action. In the complex field of dementia care, understanding what strategies work (6) is key to improve the quality of care for people living with dementia.

One approach widely used to implement evidence-based practices is the quality improvement collaborative (QIC) (7). This approach developed by the Institute for Healthcare Improvement (8) involves bringing together health professionals to learn and share methods to improve care. Key elements include a focus on a specific topic of healthcare, participants from multiple sites, clinical and quality improvement experts to provide advice and guidance to participants, structured activities to identify and try out improvements over time, and monitoring of progress against the aims of the improvement (9, 10). Despite their appeal in improving healthcare, high set up costs and varied success limit confidence in their use (7, 11, 12). There are few examples of QIC applications in dementia care (13–15).

Recent studies of QICs have described components (9, 16), reported on evaluations (17, 18), effectiveness (7) and cost-effectiveness (12), and identified factors influencing outcomes (19–21). Researchers identified the need to open the ‘black box’ of QICs to understand how components contribute to success (10, 22, 23). A theory-based understanding of the QIC process is advocated to better understand the influence of context on outcomes (24, 25). Understanding how and why QICs work under different circumstances is critical to assess suitability and justify the approach. Complex interventions such as QICs are multicomponent processes that interact with each other and the external and organisational contexts in which they operate (26). Linking a theoretical approach to an evaluation framework helps better understand how to design implementation interventions and evaluate how they work. Realist approaches have been used to understand how QICs work (25) and several studies have reported realist evaluations of process and outcomes (27–29). Few studies have used a realist approach (30) or explored the use of QICs to improve quality of dementia care (31). Realist evaluation (32) provides a method to understand how clinicians build knowledge and skills to improve dementia care in diverse settings (33).

## Methods

### Aim

This realist-informed process evaluation aims to improve our understanding of the ways in which QICs work to build knowledge and skills in quality improvement and implementation of evidence-based guidelines into practice in dementia care. Research questions are:

1. How, why and under what circumstances do QICs build knowledge and skills in clinicians to improve quality and practice?
2. Was the QIC approach acceptable and feasible to clinicians?

The process evaluation was embedded within a translational research trial (referred to as ‘Agents of Change’) which examined whether QICs could improve adherence to several recommendations in the Australian Clinical Practice Guidelines and Principles of Care for People with Dementia (referred to as the guidelines hereafter) (34). Full methods for the trial have been published in a protocol paper (34). The effect of the QIC on the outcome of guideline adherence was measured using an interrupted time series design and results will be reported separately. Ethical approval for the study was granted by the Southern Adelaide Clinical Human Research Ethics Committee (HREC/17/SAC/88). Clinicians responded to advertisements for the collaborative and self-selected to join a sub-group within the collaborative related to exercise, carer support or occupational therapy recommendations. A light touch, low cost intervention was trialled. This included online learning modules, teleconference meetings and email communication to reduce time and costs of participation. Local adaptation of the guideline recommendations was encouraged.

### Patient and Public Involvement

Experts by experience of dementia (people with dementia and care-partners) were involved throughout the Agents of Change trial. The roles included membership of the Investigator and management group (JT), identification of priorities for the trial, review of training modules for collaborative topics, presentations to participants, and feedback on implementation plans. They were not involved in the design of this study. JT as co-author reviewed the manuscript and offered comments and wording for acknowledgement. An evaluation of the impact of experts by experience of dementia will be reported separately. Their contribution is acknowledged at the end of this article. Supplementary file 1 summarises the components in the QIC.

## Study Design

This evaluation followed available guidance on process evaluation (35, 36) and realist evaluation (32) in knowledge translation interventions (37). It addressed implementation outcomes of acceptability and feasibility of the QIC approach in building skills and knowledge of participating clinicians (27). Outcomes of fidelity, penetration, and uptake of the clinical guidelines for dementia care as described in the protocol paper are addressed elsewhere (34).

The study was completed in four phases:

Phase 1: Development of the initial program logic and program theory to be tested This involved: 1) describing the strategy and logic of the program, 2) considering the context (C) of the intervention, 3) identifying the underlying mechanisms (M) and 4) reporting on the implementation outcomes (O) achieved (35). This is denoted as Context (C), Mechanism (M) and Outcome (O) configurations (4, 16) to enable understanding of the relationship between these program aspects.

The initial program theory was developed through searches of grey literature for theory components and academic database searches (26). The multiple components of the QIC method (38) were explored, then theory components were identified (39). This was confirmed with stakeholders using ‘If…then’ statements (Figure 1-part A), to be tested with clinicians.

Phase 2: Pre-and post-intervention data collection of surveys and interviews (quant +QUAL)

A concurrent mixed methods approach (40) was used to develop an understanding of the clinicians’ experience in the QIC. The survey tool QIKAT-R (41) identified changes in their level of knowledge of quality improvement over two time points. The NoMAD survey (42) was used to identify clinicians’ understanding of processes to normalise changes in practice on commencement and at completion of the program. These measures were compared with the mechanisms of change identified by clinicians in interviews, to examine patterns hypothesised in the initial program theory (39).

Phase 3: Data analysis, outcome patterns, and hypothesis testing

Survey and interview data identified change in knowledge and skills, and clinicians’ experience of the collaborative. Exit interviews with clinicians who withdrew from the collaborative provided an understanding of their decision. The reasons for how and why the collaborative process worked for clinicians was configured as context (C) + mechanism (M) = outcome (O) for each component. Summaries of the configurations were presented for three major settings in which clinicians worked. The interviews described acceptability and feasibility of the QIC and how learning generated change (43).

Phase 4: Refinement of the initial program theory

Survey data and identified configurations were tested against the initial program theory to confirm, refute, or revise the theory of how and why the QIC worked and in what circumstances (39). Where patterns matched hypotheses the program theory was confirmed. Where data did not match, the hypothesis was refuted and where additional conditions were identified, the program theory was modified. A revised program theory was developed to understand how QICs build knowledge and skills for clinicians (37)

## Data Collection

### Surveys

Quantitative data were collected in two surveys, using QIKAT-R (41) and NoMAD (42) administered pre-and post-participation in the collaborative. QIKAT-R is designed to assess clinicians’ ability to write an aim, a measure and change for a quality improvement scenario (41). NoMAD (42) assessed the degree of agreement of clinicians with statements based on the four Normalization Process Theory (NPT) (44) constructs of normalising a change to practice.

Clinicians consented to participate in the evaluation and undertook the surveys online in the introductory and final learning modules. Data were extracted for analysis in the evaluation. Supplementary file 2 provides an outline of the interview questions and Supplementary files 3 and 4 provide an example of the NoMAD and QIKAT-R surveys used in the online learning modules. On completion of the program clinicians were asked to comment online on their degree of success in implementing change.

### Interviews

Clinicians were invited to participate in interviews and were introduced to the evaluator via an email from the project coordinator (MCa). The first author (LdlP) undertook the evaluation as a PhD student with experience as a clinician in aged care and sought consent via the approved ethics process.

Semi-structured private telephone interviews, up to an hour, were conducted with clinicians, on commencement and completion of the program. The same questions were asked of each person to describe their motivations, experiences, setting and role. A realist interviewing approach was used to understand their reasoning and responses (45). Interviews were recorded and transcribed with consent, checked for accuracy, and sent to clinicians for comment or addition. Field notes made by LdlP during the interviews added information for accuracy, emphasis, or requests to stop recording of parts of the interview.

## Data Analysis

### Surveys

Responses were extracted from the online surveys, de-identified for each clinician and compared to identify change in knowledge and skills of quality improvement (QIKAT-R), and engagement in processes of normalising implementation (NoMAD). Results were scored (by LdlP and GR) for QIKAT-R (41) using the rubric provided. The principal researcher (KL) resolved any discrepancies. The NoMAD (42) survey responses were converted to a five-point Likert scale (46) (by LdlP and checked by GR). Descriptive statistics were used to present the spread of responses from clinicians. Small sample sizes and lack of controls limited further statistical analysis.

### Interviews

Interview data were transcribed verbatim, de-identified and entered into NVivo-12 software (47), for analysis using a combined inductive and deductive (48) framework analysis approach (49). Three frameworks were used to identify: issues related to the context (CFIR) (50), the social processes involved in normalising the change (NPT) (44), and the mechanisms at work within the collaboratives and the broader context (RE) (32). These frameworks aligned to focus on context (C), mechanism (M), and outcomes (O) to understand how, why and in what circumstances the collaboratives may work. Table 1. shows the alignment of these frameworks.

**Table 1.**
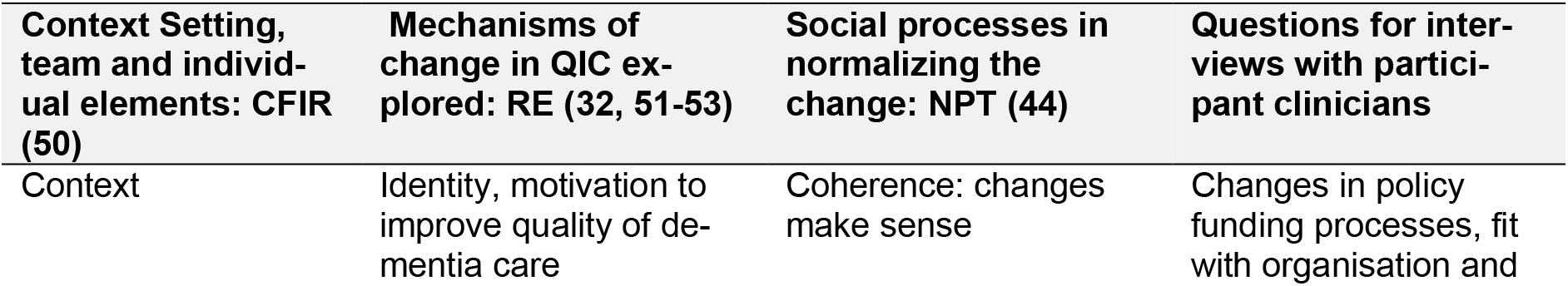
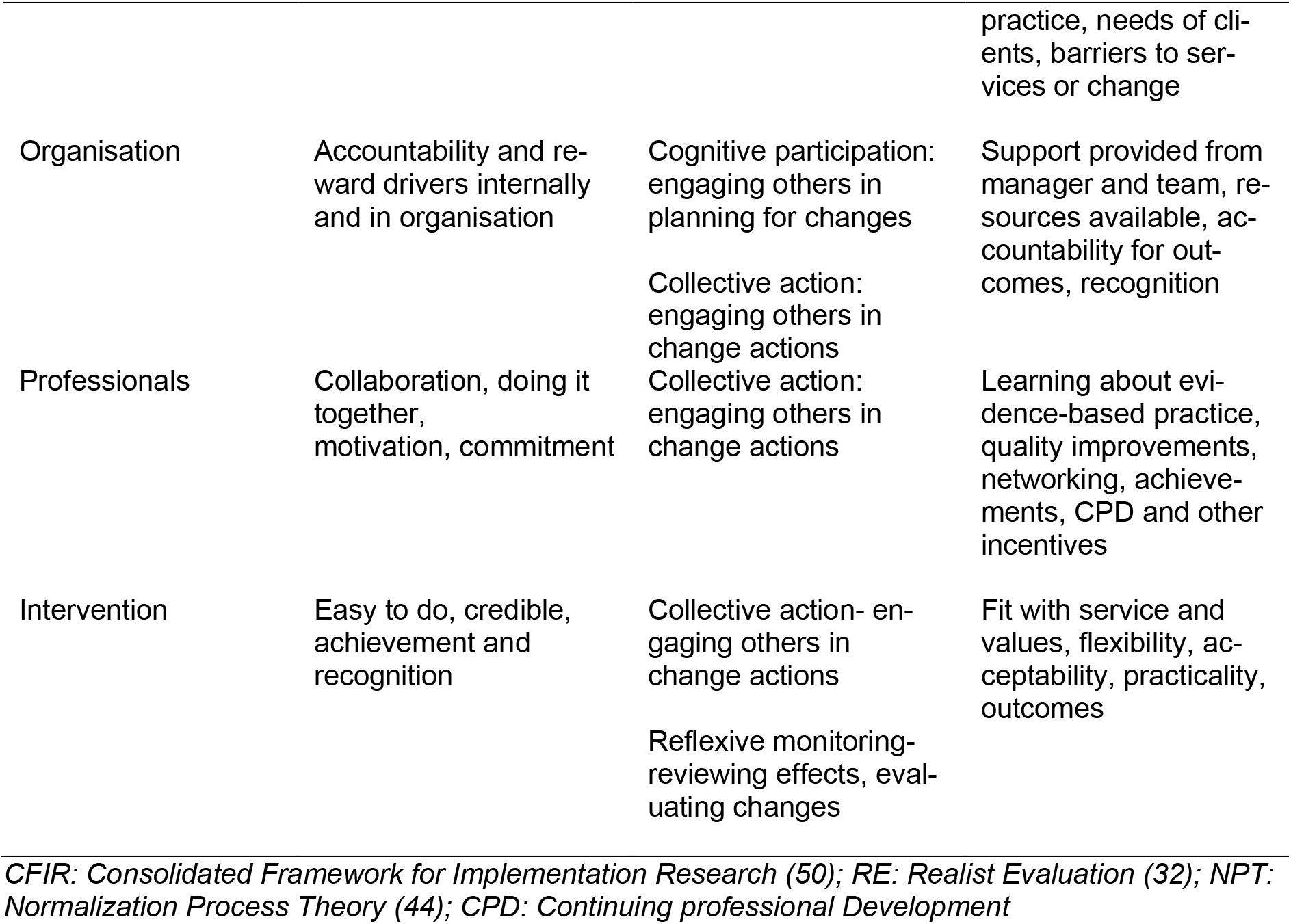
Alignment of frameworks for analysis of qualitative data.

Coding categories were developed from the frameworks and interviews were coded deductively (LdlP) with 30% checked for consistency (GR). Any differences were resolved by discussion or consultation with the principal researcher (KL). Elements of context, mechanism and outcomes patterns were searched for in the data through a deliberate inductive process (27). Quotes from interviews were extracted and presented in the results. This adapted framework analysis was used to refine, confirm, or refute initial program theory (54).

### Integration of results

Data from interviews and surveys were integrated at both the pre-and post-intervention stages through description and joint display (55) to identify where they confirmed, refuted, or modified the initial program theory. A revised program theory was developed to explain how and why the collaborative built knowledge and skills in quality improvement.

## Results

### Participants

Of the 45 clinicians in the Agents of Change trial, 28 (62%) were involved in the process evaluation. The QIKAT-R was completed by 26 (58%) clinicians at pre-intervention and 18 (38%) at post-intervention. The NoMAD survey was completed by 13 (29%) clinicians at pre-intervention and 15 (33%) post-intervention. Table 2 presents the characteristics of clinicians, showing the range of professions, settings, locations, type of organisation as well as the subgroup chosen for the collaborative.

**Table 2.** Characteristics of participant clinicians by collaborative sub-group in the process evaluation

| Collaborative sub-group | n (%) |  |  |
| --- | --- | --- | --- |
|  | Exercise<br>n=12 | Carer support<br>n=6 | Occupational<br>Therapy<br>n=10 |
| Female | 10 (83%) | 6 (100%) | 10 (100%) |
| Male | 2 (17%) |  |  |
| Regional/rural/remote | 3 (25%) | 2 (33%) | 2 (20%) |
| Profession |  |  |  |
| Physiotherapy | 10 (83.4%) | 1 (16.7%) | 0 (0%) |
| Occupational Therapy | 0 (0%) | 1 (16.7%) | 10 (100%) |
| Nursing | 1 (8.3%) | 2 (33.3%) | 0 (0%) |
| Medicine | 1 (8.3%) | 0 (0%) | 0 (0%) |
| Dietetics | 0 (0%) | 1 (16.7%) | 0 (0%) |
| Health services | 0 (0%) | 1 (16.7%) | 0 (0%) |
| Organisation Type |  |  |  |
| Public | 3 (25%) | 3 (50%) | 4 (40%) |
| Private | 2 (16.7%) | 0 (0%) | 0 (0%) |
| Not for profit | 7 (58.3%) | 2 (33.3%) | 4 (40%) |
| Sole provider | 0 (0%) | 1 (16.7%) | 2 (20%) |
| Service setting |  |  |  |
| Acute | 1 (8.3%) | 1 (16.7%) | 3 (30%) |
| Sub-acute / Transition Care | 2 (16.7%) | 0 (0%) | 1 (10%) |
| Community / Outpatient | 2 (16.7%) | 5 (83.3%) | 6 (60%) |
| Residential | 5 (41.6%) | 0 (0%) | 0 (0%) |
| Residential and Community | 2 (16.7%) | 0 (0%) | 0 (0%) |

### Pre-intervention survey results

Results for surveys are presented in Supplementary file 5 (NoMAD) and Supplementary file 6 (QIKAT-R) in comparison to post-intervention results. Most clinicians (80%) scored poorly on the QIKAT-R prior to the intervention, demonstrating limited knowledge about quality improvement. This finding validated the need for learning.

In the NoMAD survey, over 70% of clinicians saw the need for change, and how the guidelines differed from their current practice. Over 70% valued the proposed changes and 90% were optimistic about the support they would have from managers. Over 50% thought resources and training would be sufficient but only 40% were confident in their co-worker abilities to implement the changes.

### Pre-intervention interview results

Interviews were conducted with 24 (53%) clinicians. They reported feeling highly motivated to undertake the process and participate in the collaborative sub-groups. Over 85% reported having no experience of leading quality improvement processes and were unsure of their knowledge or how the implementation process would unfold in their setting.

Context:

Most clinicians identified external policy and funding constraints on their organisations which could impact on their practice. This was reflected in changes to their roles, restructuring of the organisation and time constraints.

> *“We’re going through a major…change with the new CEO…challenge for me is that because staff are unhappy, we are having a high turnover^1^’* (participant S11)

In government funded hospital settings, multidisciplinary teams and formalised quality improvement structures were identified as being supportive of the proposed changes. In aged care settings, however, most participants identified role boundaries and workloads as barriers to quality improvement.

> *“No doubt there’ll be a bit of resistance from … staff, … ‘why should I do it your way, I’ve done it this*
>
> *way my… entire life?’”* (participant E11)

For clinicians working part time, any quality improvement activity had to be done in their own time. Sole providers expected to work in their own time to improve practice but sought support from their networks and peers.

> *“it will take a little bit of time …and work outside of those hours of working face to face with them - I’m*
>
> *prepared to do that”* (participant C05)

Wait and see approach:

Most clinicians were unaware of the existence of the guidelines before commencement and were uncertain of how recommendations could be adapted to their practice. While most understood how the QIC would work, they were cautious about what would be required in their setting.

> *“I need to have a little bit more understanding of what will be required from me… to see how my work practice … fits in with what everybody’s looking for”* (participant O09)

Implementation processes:

Clinicians in all settings understood the intent to adapt implementation of the guidelines to suit their setting and expressed confidence that this approach was acceptable and feasible.

> *“I feel fairly confident that we will be able to get things off the ground and make some changes”*

(participant S15)

Those working in hospital settings were more likely to have experience of quality improvement and had begun to identify who they needed to involve in the change process.

> *“.needs to go through my director and the …reference group …so any reporting back on any changes in process or procedure …would have to be verified … and approved’* (participant C06)

Most clinicians understood that implementing changes would involve communicating with others and engaging them in new practices.

Mechanisms:

The mechanisms identified in interviews were similar but described differently by clinicians in each of the three main settings. Table 3 presents mechanisms identified across three settings for participants.

**Table 3.** Mechanisms identified across three key settings, with example quotations

|  | Public Hospital services n=14 | Residential and Community Aged Care n=6 | Private practitioners n=4 |
| --- | --- | --- | --- |
| **Motivation and building confidence to engage in change** | Improved job satisfaction and interest in dementia care were motivators<br>*"I hope to improve the service that I'm delivering and gain knowledge and confidence" E10* | Encouragement to improve services after stress of changes and interest in dementia care<br>*"if it improves the quality of life of our residents it's worth doing" E11* | Broadening business goals and interest in dementia care<br>*"it just provides us with another option that we can then promote to future clients for the business" E04* |
| **Accountability to strengthen commitment to change** | Formal staged schedule to fit in with time constraints<br>*"I think the structure of that agents for change... with how we develop a project as such, will help me" C06* | Structure to guide process and provide flexibility<br>*"...guided through and supported through the whole thing and not left to your own devices" O05* | Regular reminders to keep the collaboratives as a priority<br>*"if you're doing it on your own, it'll sort of get pushed to the back again" E08* |
| **Sense of identity reinforced** | Professional leadership in services<br>*'I think it is a really transferrable skill in demonstrating leadership and giving people opportunities to step up' S13* | Commitment to improved quality of services for people with dementia<br>*"I'm very passionate about people with dementia so with my values I want to make sure that they're maintaining their independence and participating in things they want to participate in." O04* | Specialist provider to people with dementia<br>*"I'm aiming for our OT practice to be specialist in services for older people" O03* |
| **Collective learning increases mutual support** | Sharing knowledge for improvement was valued<br>*"...breadth of the experience... from the team itself will be really valuable to share" C06* | Learning from others and comparing interventions helped assess services<br>*"...you can pool your ideas and see where the problems are, who's having success in certain areas" E05* | Sharing knowledge enhanced satisfaction in the work<br>*"Feeling confident that I'm following best practice which, for me, creates better job satisfaction" C05* |
| **Doing it together increased safety to learn and make mistakes** | Overcoming isolation and providing confidence to learn<br>*"I really just want that contact because... I was quite out of my depth, going I don't know if I'm going in the right direction" E09* | Re-energising by working with like-minded others<br>*"Great to have so many likeminded people in the one place." C08* | Practical guided approach motivated participation<br>*"I see it as being more practical, which appeals to me" O07* |
| **Credibility increases trust and acceptance of the process** | Valued evidence base and shared focus on improvement in process<br>*"I mean, the evidence is really there and it's exciting to work with people who are on that same train of thought. That's the joy of it" O09* | Evidence base fits with accreditation standards<br>*"...recognition that it's obviously a project that would be of interest to the organisation" C08* | Connection between best practice and research<br>*"...people Australia-wide who have been involved in it and are basing their practice on research and the evidence" C05* |

### Post-intervention survey results

Results for the post-intervention surveys are presented in Supplementary files 5 (NoMAD) and 6 (QIKAT-R) to compare with pre-intervention results. Clinician scores on the QIKAT-R rose modestly, with 80% showing improvement. This suggested that they had built quality improvement skills. The results from the NoMAD survey showed that most clinicians (85%) found the process made sense, and all highly valued the changes. The training and support through the QIC were considered sufficient for 75% of clinicians to develop implementation plans and make changes to their practice. While confidence in co-workers’ abilities increased, the level of support in their workplaces decreased 15% from initial expectations. All clinicians agreed that they were able to modify practice and deliver changes.

### Post-intervention interview results

On completion of the QIC, interviews were conducted with 16 (36%) clinicians. Most reported their acceptance of and satisfaction with the process. They identified significant skills and understanding gained from the process.

> *“A solid methodology and a solid quality improvement plan have been really critical in getting us to a point where it’s working and sustainable”* (participant S13)

They reported how the process enabled them to review their own practice.

> *“.quite a bit of reading and reflection that was involved in the project, especially when you’re going*
>
> *through that PDSA cycle”* (participant C05)

However, they reported that engaging others in their workplace was demanding. For those who were successful in making change, the support of managers and involvement of others were key to implementing the guidelines. Clinicians in aged care and public health settings reflected on the team effort. “*.it was really a team effort at the end of the day. “* (participant S13)

> *“……identifying your local heroes and putting responsibility on other people …, ‘this isn’t just me doing this. This is us doing this’”* (participant E11)

Others were able to align the improvements with organisational strategies and structures and gain support from others. In public health settings with quality improvement structures, this alignment made the process feasible and provided both accountability and recognition.

> *“. it crosses over many of the domains from the organisational point of view and accountability…. It’s been great to have that recognised”* (participant C06)

External and internal context changes led six clinicians (13%) to withdraw from the program. Two had personal family circumstances that led them to leave their work and participation in the program. Others were related directly to organisational changes.

Funding changes at a national level resulted in significant organisational and role changes and stress for two clinicians in aged care settings.

> *“…the sector is facing quite dramatic reform…, - our focus upon managing dementia in the community, may not be a priority going forward”* (participant S06)

That led to changes in the level of support available from their managers.

> *“…the support from management is very limited because their energy is all being focused on the (organisational) change itself.”* (participant S02)

In public healthcare, time constraints impacted on the level of inter-disciplinary team support, with one clinician withdrawing due to tensions in the team.

> *“The dynamics were more difficult than I had anticipated, and making any change was going to alienate me”* (participant O08)

Those who left the collaboratives were disappointed in not completing the program. They valued the learning modules, access to peers, and research team support. Feedback to the project team was positive with suggestions to reduce the lead-in time and the preference of having another colleague working with them for support.

Mechanisms at work within the collaborative:

The initial program theory was shared with clinicians in the interviews to consider and reflect on their experiences. The *’if…then’* propositions were presented to clinicians to assess if and how each applied to them. The mechanisms identified on commencement were generally supported and some were modified on reflection. The structured process of the collaborative provided confidence while a sense of accountability to complete the program drove commitment to the changes. The collaborative provided a sense of community and confidence in the process. The credibility of the evidence base and the team of experts and researchers engendered trust. An additional mechanism was identified of how achievements were recognised through reflection. Table 4 summarises the mechanisms and reasoning identified.

**Table 4.** Summary of mechanisms identified by clinicians at the conclusion of the program, with example quotations

|  | **Public Hospital services**<br>**n=10** | **Residential and Community Aged Care**<br>**n=4** | **Private practitioners**<br>**n=2** |
| --- | --- | --- | --- |
| **Motivation and confidence to engage in change** | *Structured and supportive process assisted engagement*<br>*"I just felt motivated throughout the process because I knew I had the support of your team, so the expertise and leadership" O13*<br>*"OTs love structure. The structure was very good, so that was great" O09* | Structured and supportive process was flexible and assisted their engagement<br>*"...it made it more appealing, and easier to engage with" O04* | Relevant and useful approach made steps practical<br>*"...could go in and do parts of it and then come back to it, that made it a lot easier to fit it in to work" C05* |
| **Accountability strengthened**<br>**Commitment to change** | *Fitted in with organisational and time requirements*<br>*"it crosses over many of the domains from the organisational point of view and accountability point of view" C06* | Maintained engagement and accreditation<br>*"I was much more motivated to do it, I felt like it had work outcomes and a personal outcome" C07* | Maintained engagement and accreditation<br>*"It was actually you guys kind of driving us to get the work done. Which is a good motivating factor for people like me who get distracted easily" E13* |
| **Sense of Identity reinforced** | Professional evidence-based practice<br>*"having an external auditor to come through and look at that and feel that it was a good project. And having great outcomes is really positive to hear too" S13* | Advocate for improved quality of services for people with dementia<br>*"...chose to work in aged care. I knew I was doing it for my residents, and to help support the staff" E11* | Professional competence in dementia<br>*"even though I've sort of worked within an aging population for a long time, I really wanted to know what best practice was" O07* |
| **Doing it together/**<br>**Collective learning**<br>**increased confidence** | Value of sharing perspectives and learning from others for improvement<br>Initial learning from others increased confidence<br>*"I think we kind of talked a lot at the beginning and then you kind of found your feet and you knew what you were doing" C01* | Overcoming isolation and gaining support<br>Motivating by working with like-minded others<br>*"It's nice to have people who I guess work - have similar mindsets and being able to bounce ideas off of them" O04* | Sense of community and overcoming isolation<br>Confidence in practice<br>*it's always good to get other people's ideas and feedback. Working in a private practice, if that's all you are doing, it can be quite isolating" O07* |
| **Credibility built trust and Confidence in process** | Trustworthy, evidence base, aligns with organisation needs<br>Research accepted by professional bodies<br>Voice of experts by experience of dementia respected<br>*"I think the value to an organisation with the quality improvement expertise, the training, the contact, the researchers and the PD development I think is extremely valuable" C06* | Evidence based<br>CPD points through a work project<br>Perspective of people with dementia useful<br>*"I think it was helpful to see that evidence of there being different types of experts" E10*<br>*'you're getting your CPD points and you're learning while you're at work, in work time' C01* | Evidence base and acceptance by professional body<br>Validity of improvements and connection with research<br>*"It was incredibly important for me" C05* |
| **Reflection on efforts helped recognise achievements** | Alignment of organisational goals and improvement in services<br>*"I think it's the thing of linking it back to the different strategic visions and values" C06* | Influencing wider service change<br>*"...take my knowledge and my actions and my words, and influence others around me" E11* | Satisfaction with competence and professional value<br>*"...that's improved my practice and sense of empowerment I guess, working with clients with dementia and their carers" C05* |

### Integration of results

Results from the post-intervention interviews and surveys were integrated and compared to the preintervention results to identify where they confirmed, refuted, or modified the program theory.

While results from the QIKAT-R survey showed modest improvement in knowledge of quality improvement methods, data from interviews provided examples from clinicians across settings that they gained knowledge and skills in quality improvement.

The results from the NOMAD survey confirmed that clinicians were engaged with the changes and made efforts to involve others in implementing changes. All clinicians agreed that feedback on implementation plans helped them to modify practice and deliver changes. The interview data confirm the value of reflective practice to clinicians to consider gaps and to monitor progress in changes.

Table 5 presents a summary of how the qualitative and quantitative results aligned to confirm, refute, or modify aspects of the program theory.

**Table 5.** Integration of main findings and alignment with program theory

|  | Interview data | NoMAD | QIKAT-R |
| --- | --- | --- | --- |
| Pre-intervention program theory | Confirmed | Confirmed | Confirmed |
| Motivation, need for learning and support | *"I'm confident in my ability so I'm hoping with the right help and guidance it will be a success"* (participant O01) | Made sense, buy in and optimistic of support | Low score on QI knowledge and skills |
| Identity and uncertainty | *"I need to have a little bit more understanding of what will be required from me before I could really go further..."* (participant O09) | Uncertainty about time |  |
| Concern about constraints and changes in setting | *"...that particular part of the sector is facing quite dramatic reform..., -our focus upon managing dementia in the community, may not be a priority going forward"* (participant S06) | Concern about team action and skills of co-workers would hinder implementation |  |
| Post-intervention Program theory | Modified | Confirmed | Modified |
| Commitment, credibility, and achievement | *"A solid methodology and a solid quality improvement plan have been really critical in getting us to a point where it's working and sustainable"* (participant S13)<br>*"...quite a bit of reading and reflection that was involved in the project, especially when you're going through that PDSA cycle"* (participant C05) | Made sense, buy in, team action and monitoring change contributed to success | Small improvement in score on QI knowledge and skills |
| Impact of context and constraint | *"...the dynamics were more difficult than I had anticipated, and making any change was going to alienate me"* (participant O08) | Lack of team action and time constraints hindered implementation |  |

QI: Quality Improvement, PDSA: Plan-Do-Study-Act cycle

A refined program theory was developed and is presented in Figure 1-part B. Support through the QIC built confidence (mechanism) for most clinicians to make changes (outcome) despite time constraints and changing funding (context). When support was lacking in their setting, those same contextual constraints led some to withdraw or only partially complete the implementation. The credibility of the experts (context) encouraged trust in the process (mechanism) by clinicians to commit to implementation (outcome). Review of the program (context) enabled reflection and recognition of efforts (mechanism) in improving dementia care (outcome).

## Discussion

A realist informed approach provided insights into how, why and under what circumstances a QIC built knowledge and skills in clinicians working in dementia care. The QIC attracted clinicians with a passion to improve dementia care in a context of resource constraints. It provided resources and opportunities for clinicians that were not usually available in their setting and met their needs for support, coaching, practice reflection and a flexible structure. They valued the credibility of the program, the flexible approach which suited their work needs, being part of a dementia specific collaborative and the process of trying out changes before adopting a new practice. When their personal motivation aligned with organisational structures and resources, clinicians successfully built the knowledge and skills to implement significant systems improvements and were recognised for their achievements. Others were able to change their practice for the selected recommendations of the guidelines and reported improvements for their clients. Many faced contextual barriers through time and resource constraints, manager or team resistance, major organisational restructures, and policy changes. While some clinicians withdrew due to contextual barriers, most gained knowledge, skills, and the confidence to engage in quality improvement which improved practice in their setting. There was a sense of empowerment for many clinicians in overcoming barriers to change. Seven mechanisms in the QIC were identified: motivation, accountability, identity, collective learning, credibility, and reflective practice. The relationships between context, mechanisms and outcomes showed how components of the QIC worked to build confidence, knowledge, and skills for most clinicians. The flexible, on-line delivery, and guidance of the QIC program made the process acceptable and feasible for most clinicians.

While QICs have been studied extensively, implementation has differed and outcomes have been inconsistent (7, 56). Few studies have used a realist approach (30) or explored the use of collaboratives to improve quality in dementia care (31). Applying a theory-based evaluation to understand how and why a QIC built knowledge and skills in clinicians, is key to capacity building (57) and identifying strategies for knowledge translation efforts internationally.

This study advances the understanding of how components of QICs contribute to success and why they matter to clinicians. It offers an understanding of a key component of collaboratives: how, why and in what circumstances clinicians build knowledge and skills in quality improvement. The findings offer insights to inform the spread of improved dementia care.

### Evaluation strengths and limitations

The use of realist-informed process evaluation was a key strength. A theory-led framework analysis offered perspectives of context, implementation process and mechanisms at work within the collaboratives. The mixed methods design offered the opportunity to gather rich qualitative and quantitative data to examine how QICs work.

A limitation of this evaluation was the use of the QIKAT-R survey to measure knowledge about quality improvement. The survey was presented in a way that led to participants focussing on clinical responses rather than a process improvement approach, resulting in low scores. The interview data provided stronger evidence of improved knowledge and skills. Small numbers of participants in the evaluation limited statistical analysis but still offered a rich exploration of the mechanisms and contextual factors affecting their learning.

## Conclusion

This study addresses a strategy to improve dementia care. A QIC designed to suit geographically dispersed clinicians in different settings and roles was acceptable and feasible in building knowledge and skills to improve dementia care. The motivations of clinicians and the credibility of the collaborative process built commitment and trust to learn implementation processes and empower clinicians to improve dementia care. This offers insight in how collaborative improvement processes work for designing QICs in complex and resource constrained settings.

## Data Availability

Consent for making individual data available was not sought. aggregated and de-identified data may be available on reasonable request.

## Acknowledgements

We gratefully acknowledge the involvement of the following people living with dementia and care-partners (Experts by Experience of Dementia) in the Agents of Change Quality Improvement

Collaborative project: Jane Thompson, Ian Gladstone, John Quinn, Glenys Petrie, Gary Collins, Mae Collins and Nadine Hedger.

